# Automation Bias in Large Language Model Assisted Diagnostic Reasoning Among AI-Trained Physicians

**DOI:** 10.1101/2025.08.23.25334280

**Authors:** Ihsan Ayyub Qazi, Ayesha Ali, Asad Ullah Khawaja, Muhammad Junaid Akhtar, Ali Zafar Sheikh, Muhammad Hamad Alizai

## Abstract

**Importance:** Large language models (LLMs) show promise for improving clinical reasoning, but they also risk inducing automation bias, an over-reliance that can degrade diagnostic accuracy.

Whether AI-trained physicians are vulnerable to this bias when LLM use is voluntary remains unknown.

**Objective:** To determine whether exposure to erroneous LLM recommendations degrades AI-trained physicians’ diagnostic performance compared to error-free AI advice.

**Design:** A single-blind randomized clinical trial was conducted from June 20 to August 15, 2025.

**Setting:** Physicians were recruited from multiple medical institutions in Pakistan, participating through in-person or remote video conferencing.

**Participants:** Physicians registered with the Pakistan Medical and Dental Council with MBBS degrees, who had completed a 20-hour AI-literacy training covering LLM capabilities, prompt engineering, and critical evaluation of AI output.

**Intervention:** Participants were randomized 1:1 to diagnose 6 clinical vignettes in 75 minutes. The control group received unmodified ChatGPT-4o’s diagnostic recommendations; the treatment group’s recommendations contained deliberate errors in 3 of 6 vignettes. Physicians could voluntarily consult offered ChatGPT-4o recommendations alongside conventional diagnostic resources based on their clinical judgment.

**Main Outcomes and Measures:** Primary outcome was the diagnostic reasoning accuracy (percentage), assessed by three blinded physicians using an expert-validated rubric to evaluate: differential diagnosis accuracy, appropriateness of supporting and opposing evidence, and quality of recommended diagnostic steps. Secondary outcome was the top-choice diagnosis accuracy.

**Results:** Forty-four physicians (22 treatment, 22 control) participated. Physicians receiving error-free recommendations achieved mean (SD) diagnostic accuracy of 84.9% (19.7%), whereas those exposed to flawed recommendations scored 73.3% (30.5%), resulting in an adjusted mean difference of -14.0 percentage points (95% CI: -8.3 to -19.7; *P* <.0001). Top-choice diagnosis accuracy per case was 76.1% (42.5) in the treatment group and 90.5% (28.9) in the control group, with an adjusted difference of -18.3 percentage points (95% CI, -26.6 to -10.0; *P* <.0001).

**Conclusions and Relevance:** This trial demonstrates that erroneous LLM recommendations significantly degrade physicians’ diagnostic performance by inducing automation bias, even in AI-trained physicians. Voluntary deference to flawed AI output highlights critical patient safety risk, necessitating robust safeguards to ensure human oversight before widespread clinical deployment.

**Trial Registration:** ClinicalTrials.gov Identifier: NCT06963957

## Introduction

Diagnostic errors remain a significant source of preventable harm globally, contributing to 5.7-8.4 million excess deaths annually in low- and middle-income countries (LMICs) and an estimated 795,000 deaths or cases of permanent disability in the United States.^1-5^ Most diagnostic errors arise from judgment-related pitfalls: physicians anchoring on narrow differentials, misinterpreting test results, or delaying specialist consultations.^6-8^ While large language models (LLMs), such as ChatGPT-4o, hold promise for reducing diagnostic errors by augmenting clinical reasoning, ^9-13^ their propensity to “hallucinate,” generate plausible but false information, poses significant safety risks.^14-16^ The extent of these errors depends heavily on how LLMs are prompted. For instance, when leading LLMs were tested with physician-validated vignettes containing even one incorrect detail, hallucination rates reached 50-82%.^16^ These risks are amplified by automation bias, the tendency to over-rely on automated output, leading clinicians to accept erroneous recommendations without adequate scrutiny.^17-21^

While these hallucination risks are concerning, the unique characteristics of LLMs may lead to novel patterns of automation bias compared to traditional AI models, such as convolutional neural networks (CNNs). Unlike traditional AI systems that provide discrete classifications with confidence scores (e.g., “malignant” with “92% probability”), LLMs generate narrative recommendations that appear highly sophisticated, yet may contain subtle but clinically significant errors. Prior studies using traditional AI models have documented the effects of cognitive biases,^22-27^ showing that inaccurate predictions impair radiologist performance and erroneous suggestions prompt pathology experts to overturn correct diagnoses in 7% of cases.^22,23^ However, LLMs’ narrative sophistication may either amplify automation bias by making erroneous recommendations more persuasive, or conversely, may reduce bias by engaging physicians in deeper analytical thinking.

Evidence suggests that physicians are willing to adjust decisions in response to LLM feedback; in one study, diagnostic accuracy improved by 18% when physicians reviewed GPT-4’s suggestions after their initial assessment.^25^ While practical, such pre-post designs fix the consultation order (AI-first or clinician-first) and mandate AI review or provide continuous AI display, thereby limiting insight into clinicians’ discretionary use of AI.^17,22-25^ In contrast, on-demand consultation models are being increasingly adopted, where clinicians consult LLMs at their discretion, as these preserve clinical autonomy, integrate naturally into existing workflows, and allow selective AI engagement based on case complexity and clinician judgment.^10-12,28-30^ Given the growing emphasis by healthcare organizations on AI-literacy for mitigating automation bias,^31-33^ a critical question emerges: Are AI-literate physicians exercising voluntary consultation vulnerable to automation bias when LLM recommendations contain errors? Randomized clinical trials addressing this question are lacking.

To address this gap, we conducted a randomized clinical trial to quantify the magnitude and patterns of automation bias among 44 physicians who completed a comprehensive 20-hour AI-literacy training program before randomization. Post-training, physicians were randomized to receive either unmodified ChatGPT-4o diagnostic suggestions (control group) or recommendations containing deliberate, clinically significant errors in 3 of 6 clinical vignettes (treatment group), with error placement randomized to prevent pattern recognition. Importantly, all physicians maintained complete autonomy to consult, modify, or ignore the LLM suggestions entirely while retaining access to conventional diagnostic resources such as online medical databases and search engines (without AI features). This design isolates the causal effect of erroneous LLM recommendations on physicians’ diagnostic performance under conditions that mirror real-world voluntary adoption, generating essential evidence for informing evidence-based guardrails for safe clinical deployment.

## Methods

Our study was approved by the Lahore University of Management Sciences (LUMS) institutional review board, and all participants provided informed consent prior to enrollment. Participants received USD 20 as compensation for participating in the study. The trial was prospectively registered on ClinicalTrials.gov (NCT06963957; first posted April 30, 2025) before participant enrollment. The study design and reporting adhere to the Consolidated Standards of Reporting Trials (CONSORT) 2025 guidelines; the complete study protocol is in Supplement 1.

Physicians with varying specialties and clinical experience were recruited through email distribution lists at the LUMS Learning Institute, which offers specialized training programs for physicians on healthcare AI and data science. Eligible participants included physicians registered with the Pakistan Medical and Dental Council, held a Bachelor of Medicine, Bachelor of Surgery (MBBS) degree and had completed a 20-hour AI-literacy training (Table S7 in Supplement 2) covering LLM capabilities, prompt engineering, and strategies for critically evaluating AI-generated output. Participants were recruited from two consecutive cohorts of the AI-training program and were supervised by study coordinators in either remote sessions or at an in-person computer laboratory at LUMS. Each session lasted 85 minutes, comprising a 10-minute baseline survey followed by 75 minutes of clinical vignette assessments.

Participants were randomly assigned (1:1) to either the treatment group (n=22) or the control group (n=22) using a computer-generated randomization sequence. To prevent priming effects and ensure valid measurement of genuine, unconscious automation bias, participants were blinded to the study’s specific aims. The single-blind randomization was known only to the study administrator. Figure 1 illustrates the participant flow, with a visual representation provided in Figure S2 in Supplement 2.

**Figure 1:**
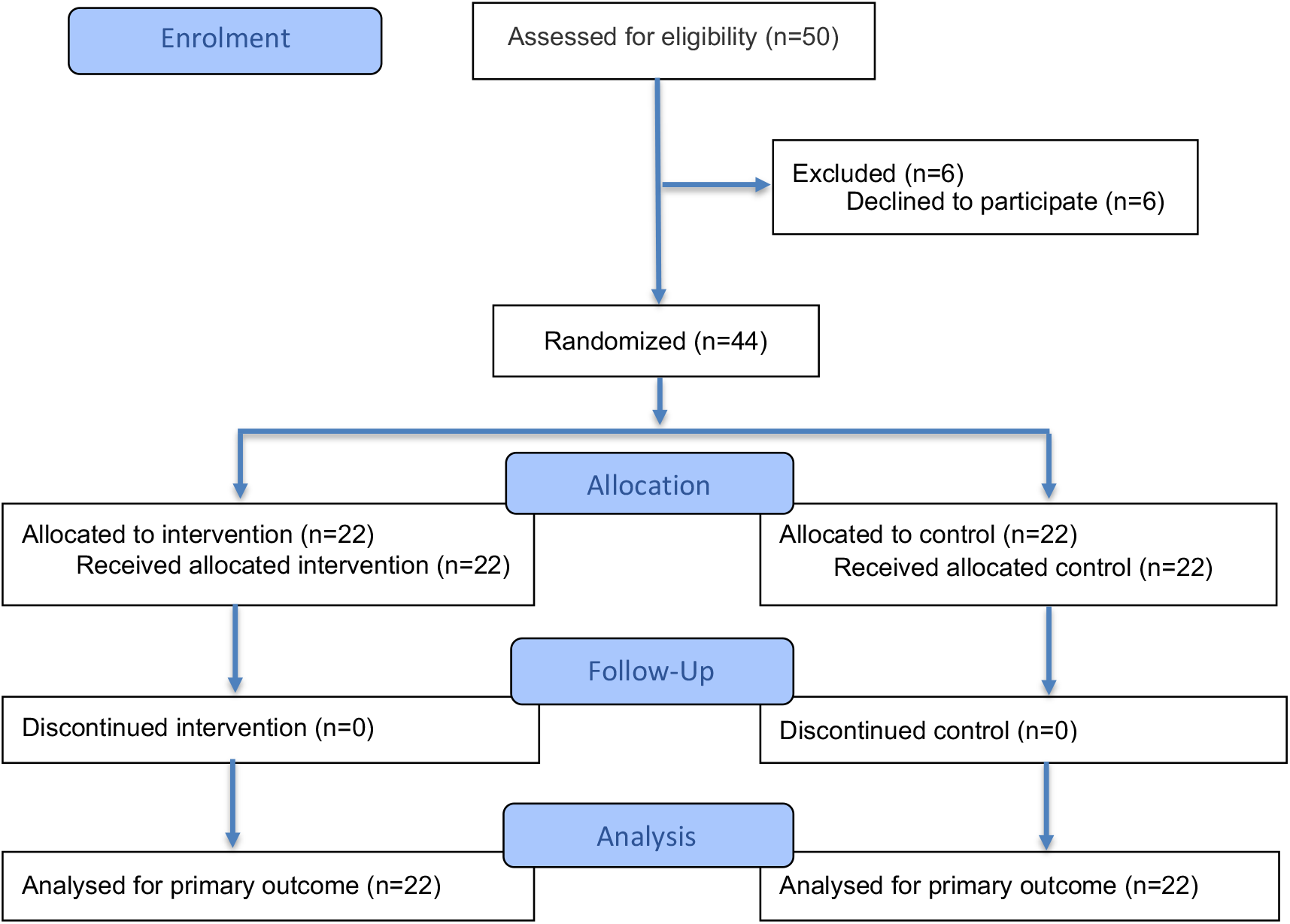
Study flow diagram. The study included 44 physicians, who completed a total 264 cases. Six expert-developed cases were presented to each physician, with scoring rubrics created by a panel of three licensed physicians w’ith expertise in clinical reasoning assessment. The control group received unmodified diagnostic suggestions from ChatGPT-4o. while the treatment gr oup received suggestions containing deliberately introduced errors for three of the six cases, which were randomly ordered to avoid anchoring bias. Physicians in both gioups could voluntarily consult the Al alongside convention diagnostic resources (e.g., PubMed. Google Search without Al features). The pre-specified primary outcome was the difference in diagnostic reasoning score between gioups on expert-developed scoring rubrics. The pre-specified secondary outcome was the most likely diagnosis per case.

**Figure 2:**
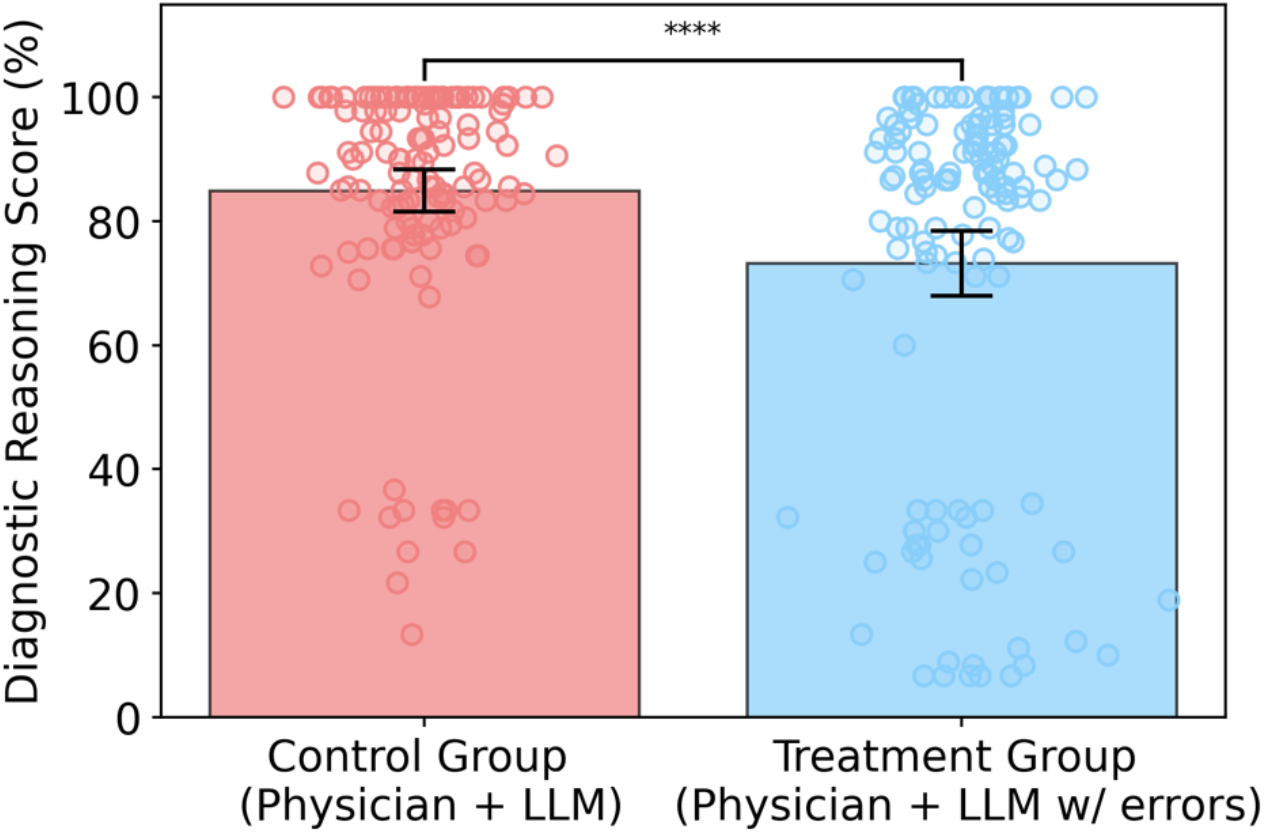
Comparison of the primary outcome for physicians in the treatment group with physicians in the control group (diagnostic reasoning score standardized to 0-100). Forty-four physicians were randomized 1:1 and completed 264 cases (132 in the treatment group, 132 in the control group). Bars represent group means with 95% confidence intervals (error bars). Individual data points show case-level scores. The treatment group demonstrated significantly lower diagnostic reasoning scores than the control group, with an adjusted difference of -14.0 percentage points (95% CI: -18.9 to -9.1; *P* <.0001) from a prespecified linear mixed effects model.

### Clinical Vignettes

Three physician co-authors (M.A.K, M.J.A., and A.Z.S.) initially developed eight clinical vignettes spanning internal medicine, cardiology, neurology, pediatrics, infectious disease, and emergency medicine. From this pool, six were chosen that offered a meaningful diagnostic challenge, excluding cases that were overly simple or rare. To ensure consistency and uniform presentation of information, all LLM-generated recommendations were pre-generated and standardized. Pilot testing established a 75-minute study time limit.

For the treatment group, the physicians embedded subtle but clinically significant errors into the ChatGPT-4o outputs for three of the six vignettes. These errors were designed to be detectable by competent physicians but not immediately apparent on casual review. The control group received error-free LLM outputs for all six vignettes. Each vignette followed a standardized format (e.g., chief complaint, history of present illness, relevant past medical history, physical examination findings, and laboratory results) to present information uniformly. A sample case is available in Table S1 of Supplement 2).

While we measured top-choice diagnostic accuracy as a secondary outcome, our primary endpoint was the diagnostic reasoning accuracy, which measured the quality of the diagnostic reasoning process. This was evaluated using structured reflection methodology that mirrored clinical practice. Using a standardized template (Table S1, Supplement 2), participants documented their top three differential diagnoses with supporting and opposing evidence, top-choice diagnosis with justification, and recommended next steps.^34^ This comprehensive assessment of the reasoning pathway, rather than just the final answer, aligns with recent literature. ^11,12^

### Assessing Diagnostic Performance

Participants’ assessment grids were scored by three physicians using a detailed rubric (Table S2 and Table S3, Supplement 2). To ensure objectivity, evaluators were blinded to group assignments, and all identifying metadata was removed from the responses. Each clinically plausible diagnosis earned up to 1 point based on its relevance and likelihood. Supporting and opposing findings identified by participants received 0-1 points per diagnosis according to correctness (0 for incorrect or missing evidence, 0.5 for partially correct or incomplete evidence, and 1 for fully correct and comprehensive evidence). The top-choice diagnosis was awarded 18 points for the most accurate diagnosis and 9 points for a plausible alternative, while incorrect diagnoses received no points. Finally, participants provided next steps to further evaluate the patient with 1 point awarded for a partially correct response and 2 points for a completely correct response based on their clinical appropriateness.

### Study Design

We employed a randomized single-blind study design. Participants were randomized 1:1 to diagnose up to six clinical vignettes in 75 minutes. Participants were randomized 1:1, with the control group receiving unmodified ChatGPT-4o recommendations and the treatment group receiving recommendations containing deliberate, clinically significant errors in three of the six vignettes, which were presented in a randomized order to prevent pattern detection. A key feature of the design was physician autonomy: consulting the AI for any vignette was a voluntary, opt-in action requiring an explicit click to view the output. Both had access to conventional online medical resources, such as medical databases and standard search, to support their diagnostic workflow. However, to isolate the intervention’s effect and prevent confounding AI exposure, a browser extension specifically blocked Google’s “AI Overviews.” Participants were instructed to approach each clinical vignette as they would in their regular clinical practice. All diagnostic assessments were collected electronically via a secure platform (Kobotoolbox). No concomitant care was applicable as participants were physicians completing assessments, not patients. Because the intervention was judged a priori to pose no more than minimal risk, we did not pre-specify individual adverse-event categories. Harms were therefore assessed non-systematically by recording withdrawals and incidents during test sessions.

### Assessment Tool Validation

To finalize the assessment rubrics, three licensed physicians with expertise in clinical reasoning assessment (M.A.K, M.J.A., and A.Z.S) independently solved each clinical vignette to flag potential scoring discrepancies. Disagreements were resolved through structured consensus discussions, resulting in standardized scoring rubrics for each case that accounted for clinical ambiguity by allowing for multiple correct variations if supported by expert consensus. Subsequently, each participant’s responses underwent blinded evaluation by three physicians. Inter-rater reliability was assessed using Krippendorff’s alpha, and the instrument’s internal consistency was measured with Cronbach’s alpha (component-wise scoring variance is detailed in Table S5, Supplement 2).

### Study Outcome

Our prespecified primary outcome was the diagnostic reasoning accuracy, calculated as a percentage of total points achieved on the assessment tool. To determine this score, three independent, blinded physicians evaluated each response using the pre-defined, validated assessment rubric, with the final score for each case being the arithmetic mean of their scores. The prespecified secondary outcome was the top-choice diagnosis accuracy, the correctness of the physician’s single most likely diagnosis for each vignette. All outcomes were compared between the randomized groups at the case-level.

### Data Analysis

We summarized outcomes with descriptive statistics. Demographic and baseline characteristics were compared between groups using χ^2^ or Fisher exact tests for categorical variables and two-sided *t* test or Mann-Whitney *U* test for the mean and median of continuous variables, respectively.

Our target sample size was 50 participants (25 per arm), based on a prior study.^9^ An a priori power analysis, conducted using Python version 3.11.9 and statsmodels version 0.14.4, indicated that 200 completed cases (4 per participant) would provide at least 80% power to detect an 8-percentage-point mean difference in scores, assuming a two-sided α of.05. The analysis employed mixed-effects models suitable for cluster-randomized designs, considering an intraclass correlation coefficient ranging from 0.05 to 0.15 and standard deviation of 16.2%. Ultimately, 44 participants enrolled and completed the study, yielding 264 completed cases (6 per participant).

Although this was 88% of our recruitment target, a post-hoc power analysis confirmed that the study remained adequately powered (≥ 80%) because the observed effect size (14.0 percentage points) substantially exceeded our initial estimates.

All analyses followed the intention-to-treat principle and were conducted at the case level, with cases clustered by participants. We used linear mixed-effects models to evaluate differences in primary and secondary outcomes. Random effects were included for participants to account for within-participant correlations and for cases to control for case difficulty variability. The secondary outcome, top choice diagnosis accuracy, was likewise without adjustment for multiple comparisons. Subgroup analyses were performed based on years of practice post-MBBS, prior LLM experience, and gender. Sensitivity analysis and robustness checks are available in Figure S1 and Table S6, Supplement 2.

All statistical analyses were conducted using Python (version 3.11.12) with the pandas library for data manipulation and statsmodels (version 0.14.4) for mixed-effects modeling. The prespecified statistical analysis plan was uploaded to ClinicalTrials.gov (NCT06963957; May 28, 2025) and is also provided in Supplement 1.

## Results

Forty-four physicians were recruited between June 20 and August 15, 2025; the participant flow is detailed in Figure 1. Of this cohort, 33 physicians (75%) attended in-person sessions while the remainder attended virtual sessions. The median (IQR) clinical experience was 10 (4.8 to 13) years. In total, participants completed 264 cases (132 per randomized group), and the trial concluded as planned. ChatGPT consultation rates were similar between the treatment (68.9%) and control (66.7%) groups (*P* =.69). Full baseline characteristics are available in Table 1.

**Table 1.**
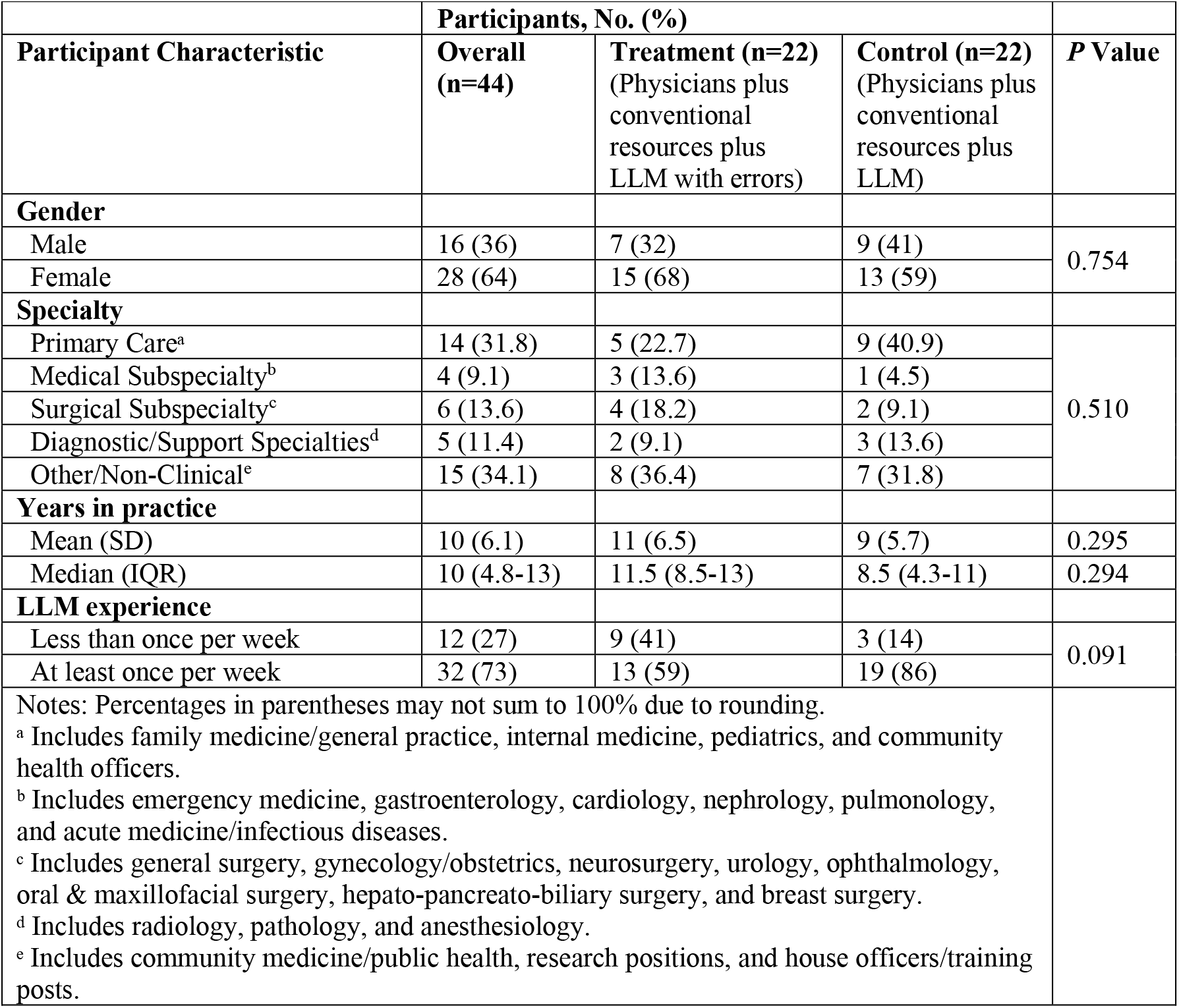
Baseline Participant Characteristics

### Primary Outcome

The control group, which received error-free LLM recommendations, achieved a mean diagnostic reasoning accuracy of 84.9% (SD = 19.7%). In contrast, the treatment group, who were offered flawed LLM recommendations in half of the cases, achieved a significantly lower mean diagnostic accuracy of 73.3% (SD = 30.5). The adjusted mean difference between the groups was -14.0 percentage points (95% CI: -18.9 to -9.1; *P* <.0001), indicating a substantial performance decline when physicians followed erroneous LLM recommendations (Table 1).

### Secondary Outcome

The mean (SD) top choice diagnosis accuracy score per case was 76.1 (42.5) in the treatment group and 90.5 (28.9) in the control group (Table 3). The linear mixed-effects model resulted in an adjusted difference of -18.3 percentage points (95% CI, -26.6 to -10.0; *P* <.0001).

### Subgroup Analyses

In prespecified subgroup analyses, we evaluated whether the treatment’s effect on diagnostic reasoning scores varied by physician experience (i.e., years of clinical practice since MBBS), self-reported use of large language models (LLMs), and gender (Table 2).

**Table 2.** Diagnostic Performance Outcomes

|  | **Mean (SD), %** |  |  |  |
| --- | --- | --- | --- | --- |
| **Group** | **Treatment**<br>(Physicians plus<br>conventional<br>resources plus<br>LLM with errors) | **Control**<br>(Physicians plus<br>conventional<br>resources plus<br>LLM) | **Difference (95% CI),**<br>**percentage points<sup>a</sup>** | **P Value** |
| **All participants** | 73.3 (30.5) | 84.9 (19.7) | -14.0 (-18.9 to -9.1) | < .0001 |
| **Years of Practice** |  |  |  |  |
| Below median | 75.1 (30.1) | 83.9 (18.9) | -9.1 (-18.1 to -0.1) | 0.0474 |
| Above median | 71.7 (31.0) | 87.0 (21.5) | -16.6 (-23.1 to -10.1) | < .0001 |
| **LLM Usage** |  |  |  |  |
| At least once per week | 71.2 (32.7) | 83.6 (20.7) | -11.0 (-18.5 to -3.6) | 0.0037 |
| Less than once per<br>week | 76.2 (27.0) | 93.4 (7.1) | -10.7 (-24.5 to 3.1) | 0.1285 |
| **Gender** |  |  |  |  |
| Male | 63.8 (36.1) | 88.6 (16.0) | -25.8 (-33.8 to -17.7) | < .0001 |
| Female | 77.6 (26.6) | 82.3 (21.7) | -2.1 (-9.8 to 5.5) | 0.5839 |
| Notes: |  |  |  |  |
| • Abbreviation: LLM, large language model<br>• <sup>a</sup> Differences between groups are reported from the linear mixed effects model accounting for clustering of cases by participants. |  |  |  |  |

**Table 3.** Top Choice Diagnostic Accuracy Score Outcomes

|  | **Mean (SD), %** |  |  |  |
| --- | --- | --- | --- | --- |
| **Group** | **Treatment**<br>(Physicians plus<br>conventional<br>resources plus<br>LLM with errors) | **Control**<br>(Physicians plus<br>conventional<br>resources plus<br>LLM) | **Difference (95% CI),**<br>**percentage points <sup>a</sup>** | **P Value** |
| **All participants** | 76.1 (42.5) | 90.5 (28.9) | -18.3 (-26.6 to -10.0) | < .0001 |
| **Years of Practice** |  |  |  |  |
| Below median | 75.8 (42.5) | 90.6 (28.7) | -14.5 (-28.0 to -1.0) | 0.0351 |
| Above median | 76.4 (42.8) | 90.5 (29.7) | -16.8 (-30.9 to -2.8) | 0.0190 |
| **LLM Usage** |  |  |  |  |
| At least once per week | 72.9 (44.6) | 89.0 (30.9) | -14.0 (-25.1 to -2.9) | 0.0134 |
| Less than once per<br>week | 80.9 (39.2) | 100.0 (0.0) | -8.1 (-27.6 to 11.4) | 0.4140 |
| **Gender** |  |  |  |  |
| Male | 61.9 (49.2) | 93.8 (23.4) | -32.8 (-46.4 to -19.2) | < .0001 |
| Female | 82.8 (37.4) | 88.2 (32.1) | -4.6 (-15.7 to 6.6) | 0.4227 |
| Notes: |  |  |  |  |
| • Abbreviation: LLM, large language model<br>• <sup>a</sup> Differences between groups are reported from the linear mixed effects model accounting for clustering of cases by participants. |  |  |  |  |

We find that the treatment effect was larger among those at or above the median years of practice (10 years), who experienced a 16.6 percentage points reduction in the diagnostic reasoning score (95% CI, -23.1 to -10.1 pp; *P* <.0001), compared to a 9.1 percentage points reduction among more experienced physicians (95% CI, -18.1 to -0.1 pp; *P* = 0.0474). The treatment effect was greater among physicians who use LLMs at least once per week, who showed a 11.0 percentage points reduction in diagnostic accuracy (95% CI, -18.5 to -3.6 pp; *P* = 0.0037), versus a 10.7 percentage point reduction among those using LLMs less than once per week (95% CI, -24.5 to 3.1 pp; *P* = 0.1285). However, the latter effect was not statistically significant at convention levels. The treatment benefit also varied significantly by gender, with male physicians experiencing a 25.8 percentage points reduction (95% CI, -33.8 to -17.7 pp; *P* <.0001) compared to a smaller and not statistically significant 2.1 percentage points reduction among female physicians (95% CI -9.8 to 5.5 pp; *P* = 0.5839).

### Assessment Tool Validation

Inter-rater reliability among three graders was high (Krippendorff’s α = 0.93), consistent with diagnostic performance studies, and internal consistency of the grading instrument was strong (Cronbach’s α = 0.80). Reliability metrics and variances for individual rubric sections are presented in Tables S4 and S5 (Supplement 2).

## Discussion

In this randomized clinical trial, we found evidence of significant automation bias among AI-trained physicians who were offered LLM’s diagnostic recommendations for clinical decision-making. Diagnostic accuracy was substantially reduced in the treatment group that received erroneous recommendations, raising important patient-safety for clinical integration. Notably, physicians were free to consult the LLM; their voluntary uptake and reliance on incorrect output indicate that technological assistance can override, rather than augment, clinical reasoning even after formal AI training. Given that AI-training is often promoted as a key safeguard against automation bias,^31-33^ our findings suggest that prior AI-training may be insufficient to offset the risk of automation bias. Safe deployment may require additional measures, such as bias-aware interfaces (e.g., provenance and uncertainty cues) and institutional oversight, alongside education.

We found notable differences across physician subgroups. Physicians with above-median clinical experience showed a greater decline in accuracy than their less experienced peers (-16.6 vs -9.1 percentage points), a finding that challenges assumptions about physician experience serving as a protective factor against AI-induced errors. This may reflect greater reliance on heuristics or overconfidence in technology among experienced physicians, which could amplify susceptibility to anchoring bias when AI provides incorrect guidance.

Gender-based differences were substantial; male physicians’ accuracy degraded significantly more than that of their female colleagues (-25.8 vs. -2.1 percentage points), with no statistically significant decline observed for the latter. This aligns with existing literature suggesting differential technology adoption patterns and verification behaviors between genders.^35,36^ Furthermore, frequent LLM users (weekly or more) showed significant performance drop (-11.0 percentage points), unlike infrequent users. This suggests that habitual AI use may foster cognitive dependency that may impair critical evaluation of AI-generated recommendations.

These analyses indicate that vulnerability to AI-induced diagnostic errors may be concentrated among experienced, male physicians with frequent LLM exposure; a demographic warranting targeted interventions.

Our findings also suggest a potential interplay between cognitive heuristics and reliance on AI. The observed automation bias may be exacerbated by factors such as cognitive offloading, where clinicians subconsciously reduce their cognitive effort when given an AI solution. Furthermore, the perceived authority and sophistication of LLMs like ChatGPT-4o might engender an unwarranted level of trust, leading to diminished critical appraisal of their recommendations.

### Limitations

Several limitations of this study warrant consideration. Clinical vignettes, while allowing controlled manipulation of variables, may not capture the complexities of real-world clinical encounters, where contextual factors, time pressures, and multimorbidity influence decision-making. Future research should explore automation bias in more ecologically valid settings. The study used deliberate errors introduced by a panel of physicians; real-world AI errors may be more subtle and harder to detect. While the participant pool was diverse in medical specialty and experience, further investigation across a wider range of healthcare professionals, including nurses and physician assistants, would be valuable. The study focused on ChatGPT-4o, chosen for its widespread commercial use, but future research should examine automation bias with other LLMs and different AI diagnostic tools. The single-session design does not address whether automation bias effects persist, diminish, or intensify with repeated AI use over time. Finally, this study did not explore mitigation strategies for automation bias.

## Conclusion

This study demonstrates significant automation bias affecting physicians’ diagnostic reasoning when using ChatGPT-4o, even when physicians are AI-trained. These findings suggest healthcare systems must implement evidence-based safeguards before widespread AI deployment, including mandatory training emphasizing critical evaluation of AI outputs and institutional protocols requiring human oversight. Future research should focus on developing interventions to mitigate automation bias, identifying high-risk physician populations and establishing frameworks for effective human-AI collaboration.

## Data Availability

De-identified participant-level data will be made available beginning three (3) months after publication in a peer-reviewed journal and continuing for five (5) years thereafter. Access will be granted to researchers associated with academic institutions. Requests should be sent to the corresponding author and will require signing a standard data-use agreement that prohibits re-identification and commercial reuse.

## Acknowledgments

We thank Ushna Malik, Muhammad Ammar Faisal, Abdullah Ghani, and Alishba Tahir for administering the surveys and proctoring study sessions.

