## Supplementary File for "Automation Bias in Large Language Model Assisted Diagnostic Reasoning Among AI-Trained Physicians" (from another version of this preprint: version 1, 2025-08-26)

**A Randomized Clinical Trial**

Ihsan Ayyub Qazi, PhD; Ayesha Ali, PhD; Asad Ullah Khawaja, MBBS; Muhammad Junaid Akhtar, MBBS; Ali Zafar Sheikh, MBBS; Muhammad Hamad Alizai, PhD

**Supplementary Content**

**eTable 1. Diagnostic Case-1 Vignette and Questions**

**eTable 2. Structured Reflection Rubric for Diagnostic Case-1 - High-Scoring Example Response**

**eTable 3. Structured Reflection Rubric for Diagnostic Case-1 – LLM Recommendation with Deliberate Flaws (Low-Scoring Example Response)**

**eTable 4. Inter-Rater Reliability by Component**

**eTable 5. Assessment Tool Validation: Component-wise Variance Between Scores Assigned by Scorers**

**eFigure 1. Statistical Validation**

**eTable 6. Sensitivity Analysis of Treatment Effects Across Model Specifications**

**eFigure 2. Study Flow Diagram**

**eTable 7. AI Training Topics**

**eTable 8: ChatGPT-4o Prompt for Diagnostic Cases**

**eFigure 3. Distribution of Diagnostic Performance Scores in the Treatment and Control Groups.**

**eFigure 4: Treatment effect by the case order**

This supplementary material has been provided by the authors to give readers additional information about their work.

### eTable 1. Diagnostic Case #1 Vignette and Questions

| Diagnostic Vignette |
|---|
| **History of Present Illness**<br>A 55-year-old female presents with a 3-month history of progressive difficulty swallowing, drooping of her eyelids, and muscle weakness. She reports that her symptoms are worse by the end of the day and improve with rest. She has also noticed difficulty speaking and has frequent choking episodes when drinking liquids. There is no associated pain, and she denies any other systemic symptoms.<br><br>**Past Medical History**<br>She has no significant medical history but was diagnosed with hypothyroidism 5 years ago and has been on levothyroxine since then. Family history is unremarkable.<br><br>**Physical Examination**<br>VITALS: BP 130/80 mmHg, pulse 75/min, temperature 98.7°F.<br>GEN: Well-appearing, no distress.<br>HEENT: Ptosis noted in both eyelids, worse on the right side.<br>PULM: Clear to auscultation bilaterally.<br>NEURO: Proximal muscle weakness<br>EXT: No edema<br><br>**Laboratory Results**<br>Acetylcholine receptor antibodies are positive. Serum creatine kinase (CK) is normal. |
| **PART 1 – Structured Diagnostic Reasoning Grid**<br>**List three possible diagnoses for the patient's condition and provide supporting and opposing evidence for each to guide differential diagnosis.** |
| **A. Diagnosis 1:** |
| a. Supporting Evidence for Diagnosis 1:<br>*Provide findings/risk factors supporting Diagnosis 1.* |
| b. Opposing Evidence for Diagnosis 1:<br>*Provide findings opposing Diagnosis 1, or findings that were expected but are not present.* |
| **B. Diagnosis 2:** |
| a. Supporting Evidence for Diagnosis 2:<br>*Provide findings/risk factors supporting Diagnosis 2.* |
| b. Opposing Evidence for Diagnosis 2:<br>*Provide findings opposing Diagnosis 2, or findings that were expected but are not present.* |
| **C. Diagnosis 3:** |

| |
|---|
| a. Supporting Evidence for Diagnosis 3<br>*Provide findings/risk factors supporting Diagnosis 3.* |
| b. Opposing Evidence for Diagnosis 3:<br>*Provide findings opposing Diagnosis 3, or findings that were expected but are not present.* |
| **PART 2 - Final Diagnostic Decision:**<br>Based upon your reasoning above, what is your final diagnosis? |
| **PART 3 - Additional Steps:**<br>**List up to 3 additional steps that you would take in your diagnostic process:** |
| Step 1 |
| Step 2 |
| Step 3 |

### eTable 2. Structured Reflection Rubric for Diagnostic Case-1 - High-Scoring Example Response

<table>
<tr><th>Question</th><th>High Scoring Example</th><th>Score</th></tr>
<tr><td>Question 1:<br>Diagnosis<br><br>List 3 Possible Diagnoses<br>(1 point each)</td><td>1. Myasthenia Gravis (MG)<br>2. Lambert-Eaton Myasthenic Syndrome (LEMS)<br>3. Hypothyroid Myopathy</td><td>3/3</td></tr>
<tr><td>Question 2:<br>Support Diagnosis<br><br>For each possible diagnosis, provide findings/risk factors supporting this hypothesis<br>(1 point each)</td><td>• Myasthenia Gravis (MG)<br>– Symptoms: Progressive muscle weakness, ptosis, difficulty swallowing, and speech problems.<br>– Pattern: Symptoms worsen at the end of the day and improve with rest, classic for MG (fatigability).<br>– Lab Results: Positive acetylcholine receptor antibodies, which are diagnostic of MG.<br>– Electromyography (EMG): Decremental response to repeated stimulation is a hallmark of MG.<br>• Lambert-Eaton Myasthenic Syndrome (LEMS)<br>– Symptoms: Muscle weakness, but typically more prominent in the proximal limbs (not as much in the neck or bulbar muscles).<br>– Pattern: Weakness may improve with activity (different from MG, where weakness worsens with activity).<br>– Lab Results: Negative acetylcholine receptor antibodies, but testing for antibodies to voltage-gated calcium channels (often positive in LEMS) would be needed.<br>– Electromyography (EMG): LEMS typically shows a facilitation response to repeated stimulation, not a decremental one.<br>• Hypothyroid Myopathy<br>– Symptoms: Muscle weakness and fatigue can occur in hypothyroidism, but ptosis and bulbar involvement are less common.<br>– History: The patient has hypothyroidism and has been on levothyroxine, though this does not explain the full constellation of symptoms.<br>– Lab Results: No significant abnormalities in thyroid function are reported, so this is less likely.<br>– Electromyography (EMG): Normal CK levels and lack of decremental response suggest this is an unlikely cause.</td><td>3/3</td></tr>
</table>

| | | |
|---|---|---|
| **Question 3**<br>**Opposing Diagnosis**<br><br>**For each possible diagnosis, provide findings opposing this hypothesis, or findings that were expected but not present** (1 point each) | • **Myasthenia Gravis**<br>– No opposing findings. The positive acetylcholine receptor antibodies, decremental response on EMG, and the symptom profile strongly support this diagnosis.<br>• **Lambert-Eaton Myasthenic Syndrome**<br>– Opposing findings: Ptosis and bulbar involvement (difficulty swallowing, choking episodes) are more suggestive of MG than LEMS, which typically has a more pronounced proximal limb weakness. The lack of facilitation on EMG also goes against LEMS.<br>• **Hypothyroid Myopathy**<br>– Opposing findings: The patient has no thyroid dysfunction on current treatment (levothyroxine). The presence of ptosis and bulbar symptoms points more toward a neuromuscular junction disorder like MG rather than a metabolic cause of myopathy. | 3/3 |
| **Question 4:**<br>**Final Diagnosis**<br>(18 points) | **Myasthenia Gravis**: Given the constellation of symptoms (ptosis, bulbar weakness, fatigue improving with rest), positive acetylcholine receptor antibodies, and decremental EMG response, the diagnosis of Myasthenia Gravis is most likely. | 18/18 |
| **Question 5:**<br>**Additional Steps**<br>(1 point each) | 1. CT scan or MRI of the chest: To rule out a thymoma, as thymic abnormalities (including thymomas) are often associated with MG.<br>2. Edrophonium test (Tensilon test): A diagnostic test that could provide further confirmation by temporarily improving muscle strength.<br>3. Assess thyroid function: Even though hypothyroidism doesn't seem to explain the full clinical picture, checking thyroid function could help rule out contributing factors or other potential diagnoses. | 3/3 |
| **Total Score** | | 30/30 |

*Caption: Example of a high-scoring response. The participant provided three plausible diagnoses with accurate and comprehensive supporting and opposing evidence and reached the correct final diagnosis.*

### eTable 3. Structured Reflection Rubric for Diagnostic Case-1 – LLM Recommendation with Deliberate Flaws (Low-Scoring Example Response)

| Question | High Scoring Example | Score |
|---|---|---|
| **Question 1:**<br>**Diagnosis**<br><br>**List 3 Possible Diagnoses** | 1. Amyotrophic Lateral Sclerosis (ALS)<br>2. Myasthenia Gravis (MG)<br>3. Multiple System Atrophy (MSA) | 1/3 |
| **Question 2:**<br>**Support Diagnosis**<br><br>**For each possible diagnosis, provide findings/risk factors supporting this hypothesis**<br>(1 point each) | • **Amyotrophic Lateral Sclerosis (ALS)**<br>– Progressive muscle weakness, especially proximal weakness.<br>– Fasciculations in the arms and legs, hyperreflexia, and a positive Babinski sign.<br>– Slurred speech, dysarthria, and ptosis.<br>– Widespread fasciculations and fibrillation potentials on electromyography, which is highly suggestive of motor neuron disease (ALS).<br>• **Myasthenia Gravis (MG)**<br>– The ptosis and difficulty swallowing suggest myasthenia gravis, but the lack of sensory symptoms and muscle atrophy make this diagnosis less likely. Acetylcholine receptor antibodies are positive, but the low titer is possibly incidental.<br>• **Multiple System Atrophy (MSA)**<br>– Muscle stiffness, dysarthria, and difficulty with fine motor tasks could be seen in MSA, but the absence of autonomic dysfunction (e.g., orthostatic hypotension) and the lack of significant MRI findings make this less likely. | 1/3 |
| **Question 3**<br>**Opposing Diagnosis**<br><br>**For each possible diagnosis, provide findings opposing this hypothesis, or findings that were expected but not present**<br>(1 point each) | • **Amyotrophic Lateral Sclerosis (ALS)**<br>– No sensory deficits, normal brain MRI and normal lumbar puncture findings argue against other causes like demyelination or autoimmune conditions.<br>• **Myasthenia Gravis (MG)**<br>– The low titer of acetylcholine receptor antibodies, lack of sensory symptoms, and muscle atrophy suggest that ALS is more likely.<br>• **Multiple System Atrophy (MSA)**<br>– No autonomic symptoms or clear MRI findings point away from MSA as the diagnosis. | 0/3 |
| **Question 4:**<br>**Final Diagnosis**<br>(18 points) | **Amyotrophic Lateral Sclerosis (ALS):** The clinical presentation of progressive muscle weakness, fasciculations, hyperreflexia, slurred speech, and motor neuron disease findings on electromyography strongly points to ALS. | 0/18 |

| | | |
|---|---|---|
| **Question 5:**<br>**Additional Steps**<br>(1 point each) | 1. Referral to a neurologist for further assessment and management of ALS.<br>2. Genetic testing for ALS to evaluate any familial predispositions.<br>3. Multidisciplinary care for symptom management, including speech therapy, physical therapy, and respiratory support. | 0/3 |
| **Total Score** | | 2/30 |

*Caption: Example of a LLM recommendation containing deliberate flaws, resulting in a low-scoring response. This was offered to participants in the treatment group.*

### eTable 4. Inter-Rater Reliability by Component

| Component | Krippendorff's Alpha |
|---|---|
| Diagnoses | 0.994 |
| Support Diagnosis | 0.993 |
| Opposing Diagnosis | 0.991 |
| Final Diagnosis | 0.996 |
| Next Steps | 0.984 |
| Total Marks | 0.927 |

### eTable 5. Assessment Tool Validation: Component-wise Variance Between Scores Assigned by Scorers

| Component | Variance |
|---|---|
| Diagnoses | 0.061 |
| Support Diagnosis | 0.066 |
| Opposing Diagnosis | 0.086 |
| Final Diagnosis | 0.133 |
| Next Steps | 0.156 |
| Total Marks | 0.067 |

**eFigure 1. Statistical Validation**

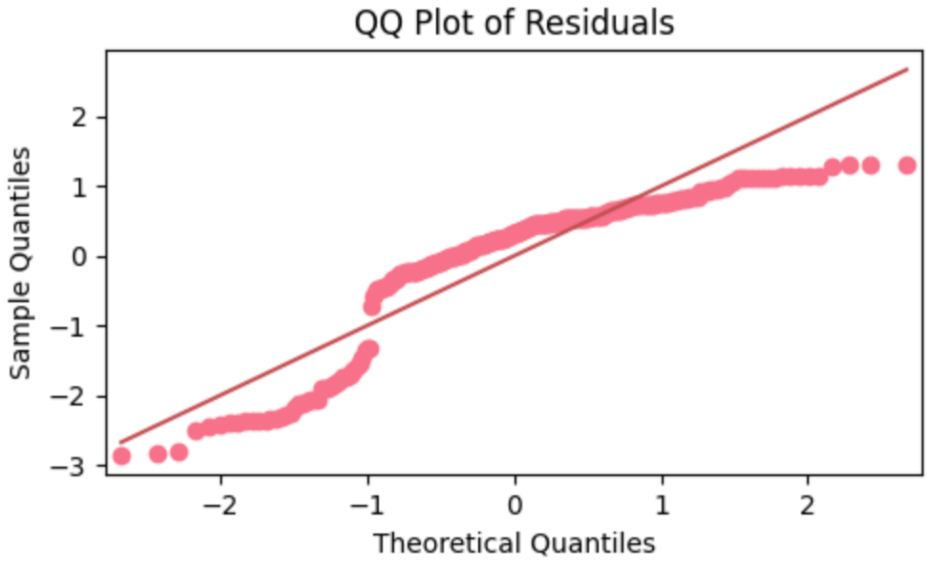


*Caption: Quantile-quantile plot of residuals from the mixed-effects model. The plot compares the distribution of model residuals (y-axis) to a theoretical normal distribution (x-axis). Points following the diagonal line indicate normality; deviations suggest departures from normal distribution. QQ plots revealed some departure from normality in the residuals, with deviations primarily in the distribution tails. However, mixed-effects models are robust to moderate violations of normality assumptions, and the observed deviations were not considered substantial enough to affect the validity of the statistical inferences.*

### eTable 6. Sensitivity Analysis of Treatment Effects Across Model Specifications

| Model | Coefficient | Standard Error | 95% CI Lower | 95% CI Upper | *P*-value | SE vs Original |
|---|---|---|---|---|---|---|
| Mixed Effects (Original) | -14.04 | 2.88 | -19.68 | -8.40 | < .0001 | 1.00 |
| Mixed Effects (Bootstrap) | -13.76 | 3.17 | -19.43 | -7.25 | < .0001 | 1.10 |
| OLS Clustered SE | -14.04 | 3.79 | -21.48 | -6.60 | .0002 | 1.32 |

*Caption: Treatment effect estimates from mixed-effects regression, bootstrap resampling, and ordinary least squares with clustered standard errors (264 completed cases, 44 participants). The coefficient represents percentage point change in diagnostic accuracy scores. Bootstrap estimates based on 500 iterations with participant-level resampling. Standard errors are expressed relative to the original mixed-effects model. All approaches show consistent evidence of significant negative treatment effects ranging from -13.76 to -14.04 percentage points, with increasing conservatism in uncertainty estimates across methods. P-values test the null hypothesis of no treatment effect using normal approximation (mixed-effects, OLS) or bootstrap distribution. Sensitivity analysis confirms the robustness of our findings to model specification and distributional assumptions.*

### eFigure 2. Study Flow Diagram

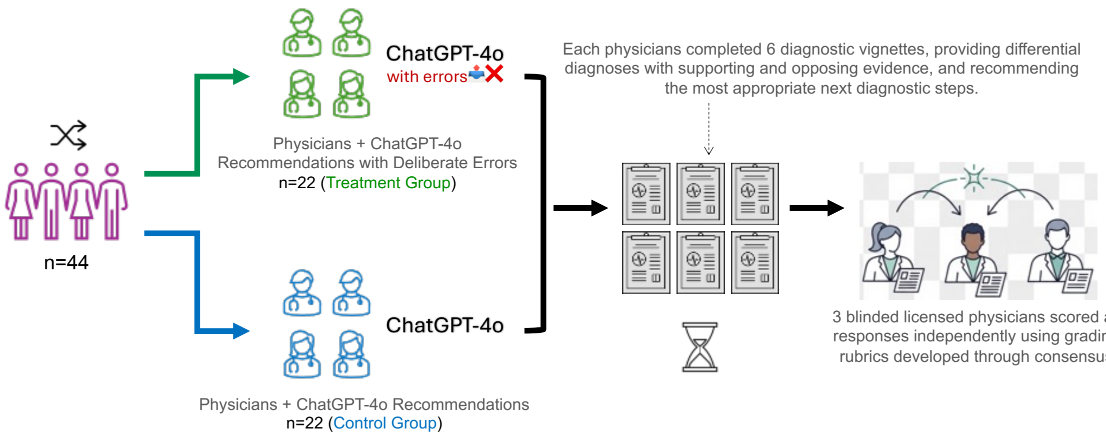


*Caption: 44 physicians completed up to 6 clinical vignettes and were randomized to either receive ChatGPT-4o recommendations containing deliberate errors (treatment group) o ChatGPT-4o recommendations without deliberate errors (control group). Participants provided differential diagnoses with supporting evidence for and against each diagnosis and recommended optimal next diagnostic evaluation steps.*

### eTable 6. AI Training Topics.

| AI Training Topics | |
|---|---|
| 1. | **Introduction to Artificial Intelligence (AI)**<br>• What is AI and why it matters<br>• AI as a helpful assistant: from mechanical tasks to intelligent support |
| 2. | **Basics of Machine Learning (ML)**<br>• What is machine learning and how it works<br>• Overview of ML types<br>• Real-life examples, including healthcare applications |
| 3. | **Basics of Generative AI (GenAI) and Prompt Engineering**<br>• How GenAI works in simple terms<br>• Introduction to prompt engineering techniques |
| 4. | **Writing and Research using GenAI**<br>• Writing professional and effective emails with AI assistance<br>• Retrieval-Augmented Generation (RAG) for improved outputs |
| 5. | **AI-powered Presentation Design**<br>• Creating presentations from descriptions and documents<br>• Prompt engineering techniques specific to presentation generation |
| 6. | **Multimedia Content Creation with GenAI**<br>• Generating and editing images using AI<br>• Creating audio and video content using simple prompts<br>• Digitizing and summarizing medical notes with AI |
| 7. | **GenAI for Data and Math Tasks**<br>• Using GenAI to explore and understand data<br>• Prompting for mathematical and data analysis tasks |
| 8. | **GenAI Customization and Limitations**<br>• Using and building your own custom GPTs<br>• Understanding AI limitations like hallucinations and how to manage them |

### eTable 7: ChatGPT-4o Prompt for Diagnostic Cases

| ChatGPT-4o Prompt |
|---|
| You are a medical doctor with an MBBS degree, who is solving a complex medical case for a test. You are going to receive a case vignette. After reading the case, I want you to give three parts of information.<br><br>Here is the first part: First, you will list three possible diagnoses. Please number these 1., 2., 3.<br><br>Next, for each of the three diagnoses listed, you will provide the findings or risk factors that support this hypothesis.<br><br>Next, for each of the three diagnoses listed, you will provide findings opposing this hypothesis, or findings that were expected but not present.<br><br>Part 2 – Final diagnostic decision. Here is the second part: Please answer this question: Based upon your reasoning above, what is your final diagnosis?<br><br>Part 3 – Additional Steps. Here is the third part: Name up to 3 additional steps you would take in your diagnostic process.<br><br>Diagnostic Case. Here is the case: |

#### eFigure 3. Distribution of Diagnostic Performance Scores in the Treatment and Control Groups.

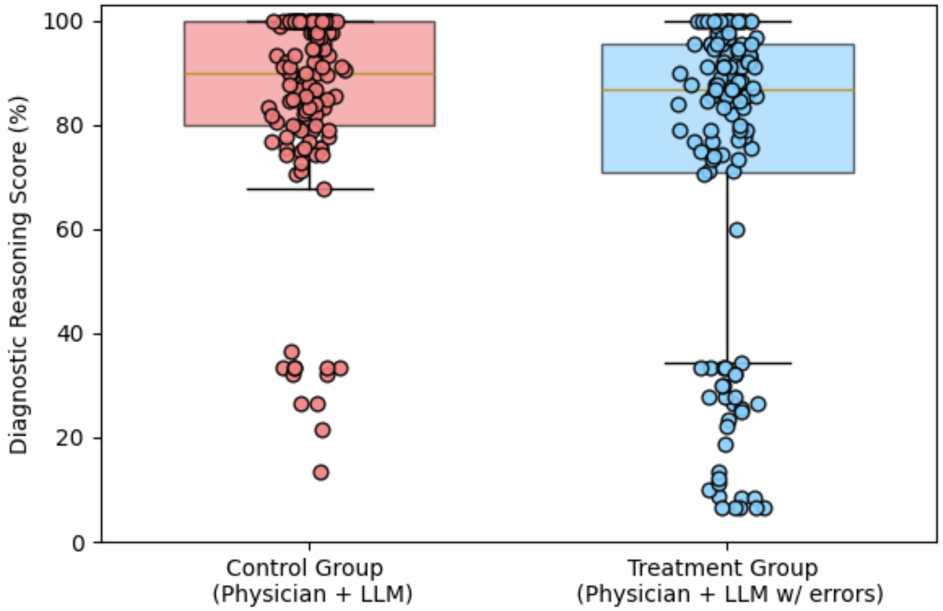


*Caption: Participants were randomized 1:1 to receive either unmodified ChatGPT-4o output (control group) or ChatGPT-4o output with deliberate clinically significant errors introduced in 3 of 6 clinical vignettes presented in randomized order (treatment group). Physicians receiving error-free advice achieved a mean (SD) diagnostic accuracy of 84.9% (19.7%), whereas those exposed to flawed advice scored 73.3% (30.5%), resulting in a adjusted mean difference of -14.0 percentage points (95% CI: -8.3 to -19.7; P < .001).*

**eFigure 4: Diagnostic score by case position order for treatment and control groups**

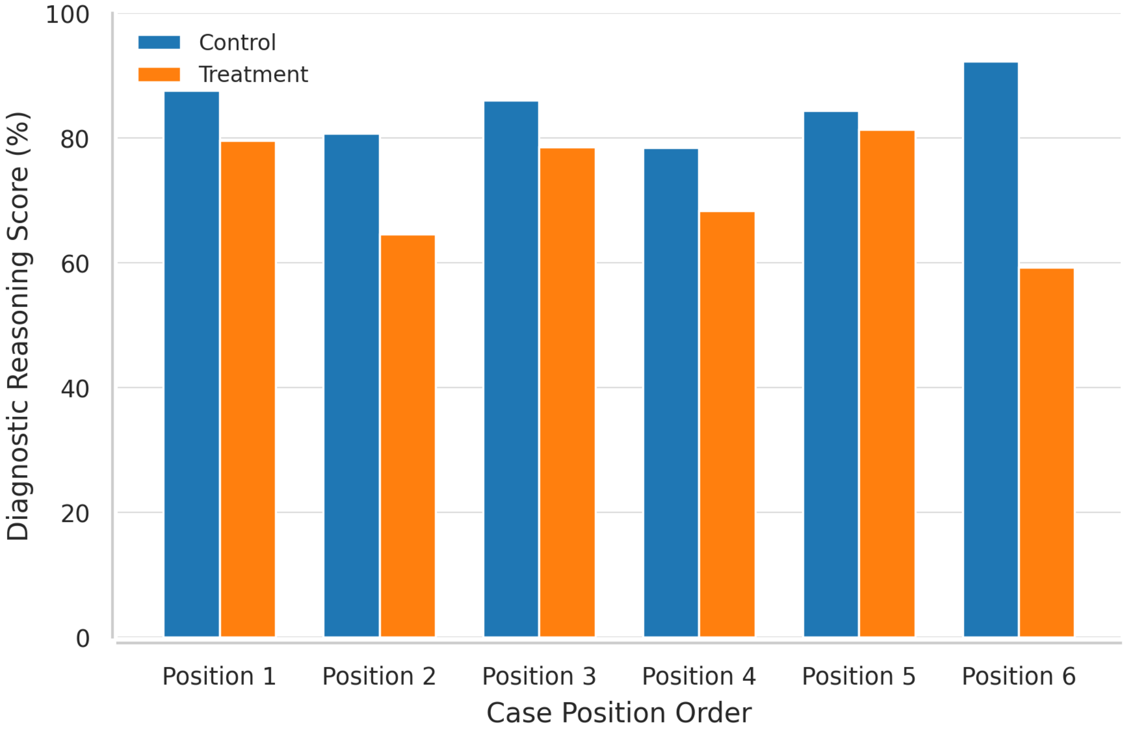


*Caption: Bar chart shows the mean diagnostic reasoning score (%) across six sequential case presentations. Blue bars represent the control group and orange bars represent the treatment group.*
